# Elucidating Post-COVID-19 manifestations in India

**DOI:** 10.1101/2021.07.06.21260115

**Authors:** Ghizal Fatima, Divyansh Bhatt, Jaserah Idrees, Bushra Khalid, Farzana Mahdi

## Abstract

**Background:** In India, a large number of patients with coronavirus disease-2019 (COVID-19), presented with common symptoms including fever, dyspnea, cough, musculoskeletal symptoms (fatigue, myalgia, joint pain) and gastrointestinal symptoms. However, information is lacking on symptoms that persist after recovery from COVID-19. In this study we assessed symptoms that persisted in patients even after their recovery and discharged from the hospital after one month from COVID-19.

**Methods:** This study is an observational cohort study. Participants in this study were enrolled between 30 to 40 days after recovery from COVID-19 of ≥18 years of age, who were hospitalized with laboratory-confirmed RT-PCR COVID-19 disease. Outcomes from post COVID-19 participants were elicited through questionnaire that consisted of three main parts beginning from subject’s demographical data, depicting about the status of COVID-19 and other co-morbidities of the subject, and about post-COVID-19 symptoms and manifestations.

**Results:** All subjects have reported some manifestation after recovery from COVID-19 whereas numerous symptoms and diseases were experienced by a great percentage of participants. Fatigue (56.25%), dyspnea (74.3%) and disturbed sleep (64.3%) were among the most common symptoms. However, more critical manifestations like renal failure and pulmonary fibrosis were reported by only a few percent of the subjects. Rating of worse physical and mental health after post-COVID recovery was also reported by subjects. There was a strong relationship found in between the presence of other co-morbidities before infection like diabetes, hypertension and in disease severity after infection. A total of 280 patients were enrolled and 160 completed the survey.

**Conclusions:** Post COVID-19 sufferers often experience symptoms that cause a disturbance in their physical health, mental health and their respiratory status for several weeks even after recovery from COVID-19. Therefore, all subjects after recovering from COVID-19 should undergo long-term monitoring programme for their symptoms and condition improvement.

## Introduction

COVID-19 has spread promptly over the past few months; however, very limited knowledge is available about long-term recovery from acute COVID-19 disease [1]. As of 26th November 2020, more than 74 million confirmed patients of COVID-19 with 1.65 million demises have been accounted worldwide. Predictions have been done that patient with only slight symptoms of COVID-19 have a major influence on cognitive, physical, psychological and also social health status. About 80% patients suffered from mild symptoms such as fever, dry cough, shortness of breath and fatigue, but in acute cases it may develop a respiratory distress or even respiratory failure and eventually increased the necessity of intensive care unit (ICU) [2].

The acuteness of the COVID-19 is associated with the age and concomitant of the diseased subjects; the older subjects were acutely disturbed and needed ICU [3]. It was also observed that the severity was also associated with the time-period of symptoms, for mild cases, the symptoms last for two weeks while for the severe cases it last for more than three to six weeks [4]. As COVID-19 is spread through exhaled air and aerosol, the major mode by which the disease transfers amid people is from the direct contact of confirmed cases of COVID-19 [5]. Polymerase Chain Reaction (PCR), Computed Tomography (CT) Scan, and blood test are the major ways of diagnosing COVID-19 [6]. However, after recovery from the disease patients have experienced other different symptoms [7]. Studies were done to understand post recovery COVID-19 symptoms, as in 2003, after the severe acute respiratory syndrome (SARS) the subjects experienced many symptoms such as myalgia, fatigue, weakness and depression. Few of these symptoms were chronic and lasted for a long time and needed long term monitoring [8].

Since the extent to which COVID-19 effects health after hospital discharge is uncertain, therefore, it is important to study the long-term consequences of COVID-19 to predict the impact of the disease, understand the natural history of the disease, and to determine the post-COVID-19 symptoms. In this study we aim to explore the post-COVID-19 symptoms, by determining the distinctive signs and symptoms the subjects experienced after recovering from the disease and these will be linked with numerous factors (age, weight, disease severity, and co-morbidities).

## Method

### Study Cohort

This is a prospective observational cohort study conducted at Era’s Lucknow Medical College and Hospital, Lucknow, India. We included post COVID-19 patients of 18 years and older with laboratory-confirmed RT-PCR test for COVID-19. We excluded patients with communication impairment and also those patients who opted out of research. This was determined by consent for this study through telephonic conversation. We used the questionnaire to assess the patient

Within 30-40 days of post covid-19. Patients who were interested agreed and registered over the call. Once well-versed approval was obtained, the study personnel surveyed (by questionnaire) the patient over the call.

Questionnaire was divided into demographic data in this we questioned the age, gender, height, smoking, and weight of the subjects, then data was collected regarding the COVID-19 level and further comorbidities of the patient like acuteness of the infection, consumption of vitamins, and other complements and occurrence of other diseases. And finally enquiry about post COVID-19 symptoms like fatigue, discomfort, shortness in breath, exhaustion etc and necessity for consuming drugs for post COVID-19 symptoms, and healing of post-COVID-19 symptoms was performed. Each feedback of subjects was analyzed to specify the degree of its existence, and then was associated with the prevalence of post-COVID-19 manifestations. Study protocol was approved by institutional ethical committee of Era’s Lucknow Medical College and Hospital.

### Statistical Analysis

The INSTAT 3.0 (Graph Pad Software, San Diego, CA) was used for statistical analysis. Quantitative variables were presented as the mean ± standard deviation. Welch’s corrected unpaired t-test was performed to assess the difference in different parameters among the mild and moderate subjects and the association of clinical characteristics among mild and moderate subjects was expressed by Fisher’s exact test which was used to obtain the odds ratio (OR). All Statistical tests were two-tailed, and p<0.05 was chosen as the level of significance.

## Results

We screened total number of 280 patients discharged COVID 19 patients, Of these 34 declined to participate, 9 found ineligible, 5 got hospitalized again and total of 232 post covid-19 patients were enrolled in the study, of whom 160 completed the survey, 107 (66.9%) male and 53 (33.1%) female. Participants were enrolled and consented to participate in the study had the mean age of 56 years, minimum age of 17 years and maximum of 88 years, length of stay of these subjects in the hospital was 10 to 11 days, they were admitted in the intensive care units. Conclusion of the reviews at a median of 40 days after the hospital discharge was done by all the partakers. From the total of 160 participants 76 (47.5%) fell under the category of mild symptoms after COVID-19 infection, 70 (43.8) fell under the category of moderate complications, and 14 participants (8.8%) fell under severe complications after COVID-19 recovery. (Table-2) Almost 90% of discharged COVID-19 patients visited or consulted doctors for their complications. 8 post discharge subjects still need oxygen after the discharge from the hospital, requiring 1-2 liters of oxygen per minute. Rest of the participants received home health aide after discharge. Other co-morbid conditions present in COVID-19 patients during their admission in the hospital are shown in table-3. Table-4 is showing the modality of COVID-19 patients and the severity during hospital admission. Treatment of COVID-19 patients during hospital admission is shown in table-5.

**Table1:** Demographic characteristic of Post COVID-19 participants:

| **Variable** | **Mean** | **SD** | **Min.** | **Max.** |
| --- | --- | --- | --- | --- |
| Age | 56.25 | 15.872 | 17 | 88 |
| Hospital stay (days) | 10.81 | 7.064 | -19 | 33 |
| BMI (Kg/m2) (Body Mass Index) | 26.9 | 4.4 | 19 | 91 |
| Health affected after COVID-19 infection | 56.85 | 36.151 | 0 | 103 |

**Table 2:** Category of COVID-19 participants:

| **COVID CATEGORY** | **Total number of participants** | **Percentage (%)** |
| --- | --- | --- |
| Mild | 76 | 47.5 |
| Moderate | 70 | 43.8 |
| Severe | 14 | 8.8 |
| Total | 160 | 100.0 |

**Table 3:** Co-morbid diseases.

| **COMORBID DISEASE** | **Total number of participants** | **Percentage (%)** |
| --- | --- | --- |
| Type 2 Diabetes Mellitus (T2DM) | 53 | 33.1 |
| Hypertension | 49 | 30.6 |
| Hypothyroidism | 6 | 3.8 |
| Hyperthyroidism | 3 | 1.9 |
| Asthma | 7 | 4.4 |
| Chronic Liver Disease (CLD) | 1 | 0.6 |
| Chronic Kidney Disease (CKD) | 5 | 3.1 |
| Tuberculosis (TB) | 2 | 1.3 |

**Table 4:** Showing the modality of COVID-19 admitted patients.

| **MODALITY** | **No.** | **Percentage (%)** |
| --- | --- | --- |
| CANULA | 2 | 1.3 |
| FACE MASK | 30 | 18.8 |
| HFNC/NIV | 4 | 2.5 |
| MECHANICAL | 1 | .6 |
| NASAL CANULA | 6 | 3.8 |
| NASAL CANULA/niv | 1 | .6 |
| NASAL PRONG | 1 | .6 |
| NIV | 2 | 1.3 |
| No | 81 | 50.6 |
| NRM | 32 | 20.0 |
| Total | 160 | 100.0 |

**Table 5:**
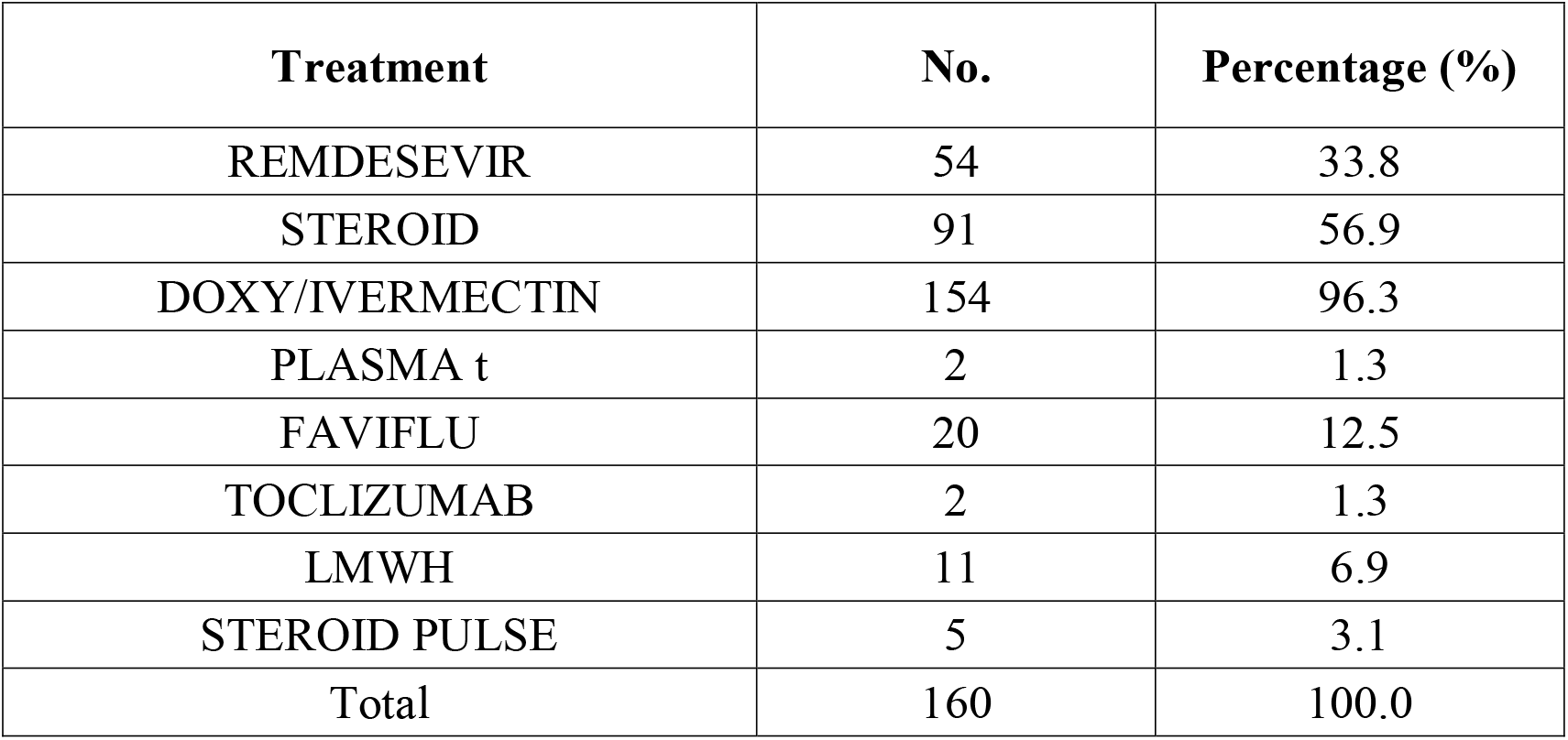
Showing the treatment of Covid-19 patients during hospital admission.

During the survey time, a total of 119 subjects reported shortness in breath, all these 119 participants during their admission in the hospital reported difficulty in breathing that doesn’t recovered even after their discharge from the hospital. In addition to this more participants reported moderate shortness in breath and severe shortness in breath during the study survey. Moreover the questionnaire revealed worst general health after infection with COVID-19 in most of the participants. Before the infection the overall health of the participants were normal without any complications, however, after the infection with COVID-19 participants lost their health, and felt more week after recovery from COVID-19. (Table-6)

**Table 6:** Symptoms of post COVID-19 patients after 40 days of recovery.

| **System** | **Condition** | **Number of participants** | **Percentage %** |
| --- | --- | --- | --- |
| Fatigue | More tired than usual | 90 | 56.25% |
|  | on daily routine | 33 | 20.6% |
| Cardiology | Chest pain | 8 | 5.0% |
|  | Palpitations | 6 | 3.8% |
|  | LL pain/swelling | 3 | 1.9% |
|  | Dyspnea | 3 | 1.9% |
|  | Orthopnea/pnd | 8 | 5.0% |
| Neurology | Anosmia | 6 | 3.8% |
|  | Agusia | 6 | 3.8% |
|  | Paraesthesia | 0 | 0.0% |
|  | Limb weakness | 30 | 18.8% |
|  | Concentration problems | 7 | 4.37% |
|  | Forgetfulness | 8 | 5.0% |
|  | Headache | 9 | 5.62% |
| Nephrology | Nausea/Vomiting | 1 | .6% |
|  | Loss of appetite | 10 | 6.25% |
|  | Decreased urine output | 1 | .6% |
| Respiratory | Breathlessness | 8 | 5.0% |
|  | Cough/sore throat/chest tightness | 36 | 22.5% |
|  | Dyspnea | 119 | 74.3% |
| Mood Disturbance | Sad | 16 | 10% |
|  | Anxious | 0 | 0.0% |
|  | Weak/tired fatigue | 120 | 75% |
| Psychiatry | Headache | 9 | 5.6% |
|  | Worry | 10 | 6.25% |
|  | Disturbed sleep | 103 | 64.3% |
|  | Hearing abnormal voices | 1 | .6% |
|  | Excess consumption of alcohol | 2 | 1.3% |

## Discussion

In our observational cohort study the Post COVID-19 patients admitted in the hospital and got recovered after being infected and discharged from hospital were surveyed for post COVID-19 manifestations, alongside a ample kind of symptoms that varied from mild symptom like fatigue and to further critical conditions like exhaustion in breath. Regarding COVID-19 toughies each one of them stated one or a few illness and upset overall health due to infection from COVID-19, and those conditions persisted even after 30 to 40 days of revival from COVID-19. However, nearly >50% patients experienced to have fatigue and exhaustion in breath for a period of more than one month since post-hospital discharge.

The intensity, frequency and duration of breathlessness and exhaustion of breath got intensified after COVID-19 in the patients who suffered from asthma and tuberculosis before the COVID-19 infection. Not only this, even both mental and physical health were graded as worst than standard of post COVID-19 infected patients following a month from the hospital discharge. In this study the acuteness of COVID-19 patients was divided in three classes, mild (less symptoms), moderate (difficulty in breathing with symptoms) and severe (breathlessness, overall health score and disturbed quality of life). The relationship in other co-morbidities present along with COVID-19 and age of participants presented a convincing association between the occurrence of other comorbidities and the acuteness of COVID-19. (9) Growing age being the most important symptom linked to severity. (10) The reported manifestations from the participants were mostly mild, so they could be reversible, like symptoms of fatigue, headache and loss in appetite. In this study it was noted that participants mostly complained of mood and respiratory problems the most, however, other problems related to nephrology (decrease in urine output) and neurology (limb weakness, forgetfulness and anosmia) were also seen and that need more attention with extra investigations requirements.

There was also a strong relationship observed in acuteness of post COVID-19 symptoms and the severity of disease during admission in the hospital, the acute cases expressed excessive disease manifestations equated with mild symptoms participants. (11) Most common manifestation for this illness was fatigue that lasts for several months (12), even in our study fatigue was present in most of the post COVID-19 patients. Another study by Lam et in 2009, followed SARS subjects for a period 4 years and evaluated the percentile of fatigue in those patients and they concluded that 40.3% suffered from chronic fatigue symptom. (13)

Further, neuropsychiatric and fatigue symptoms of large number of COVID-19 subjects were also documented by Lara et al and symptoms got worsened significantly during five weeks after the infection. (14) Forgetfulness was also reported by few subjects and those were mainly females. Effect of post COVID-19 on symptoms manifestations was recorded by Chakraborty et al. for subjects suffering from mental illness and the findings of study reported that 6% of COVID-19 subjects had worsened illness. (15) Regarding disturbed sleep in Post COVID-19 subjects it was reported that many post COVID-19 recovered subjects complained of disturbed sleep which was consistent with the findings of this study. (16) Few mild manifestations reported by participants in the study could be related to hydroxychloroquine administration like effect on vision on few subjects, receiving this drug during their course of treatment. (17) The WHO estimated the median time from commencement of disease of COVID-19 till the period of healing is about 6 weeks founded by primary data from study in China. (18) However, in India after 2 weeks or so after patient is tested negative for COVID-19 is discharged from the hospitals, while patient still have weakness and many symptoms as many participants reported shortness in breath, fatigue and disturbed sleep, with lost quality of life. Study from Italy examined post discharge patients and indicated persistent dyspnea in 43% of discharged patients’ average of 2 months after onset of symptoms, (19) in our study even higher prevalence of dyspnea was reported may be because of earlier discharge from the hospital.

Prior works on Post discharge COVID cases demonstrated that while some of pulmonary functions might retrieve, particularly in young patients, but countless others experienced reduced value of existence even after their 5 years of critical illness. (20-22) Another study by Chen et al in 2017 on long term illness outcome in Influenza A (H7N9) survivors informed improvements in respiratory functions during first six months of discharge from hospitals but displayed weakening in quality of life that continued 2 years of post discharge from hospital. (23) At this time it’s crucial to report post COVID-19 manifestations, as the survivors of COVID-19 still face so many problems by their disturbed quality of life due to fatigue and other debilitating symptoms. Furthermore, we also observed significant psychological symptoms that may disturb their capability to complete the various tasks of their day-to-day lives; therefore clinics should be set for post discharge COVID patients in order to get them proper health aide care.

## Conclusion

Most of the Post COVID-19 subjects encountered numerous symptoms even after recovery (from last negative PCR). Symptoms mostly reported by post COVID-19 subjects were fatigue, headache and disturbed sleep and more critical symptoms like dyspnea, limb weakness and other mental and physical health related symptoms. Infection severity was related to other co-morbidities present in COVID-19 infected individuals. We found that post COVID-19 participants in our study experienced many complications in regard to their health status. Therefore, subjects recovered from COVID-19 must undergo long term monitoring for their treatment of symptoms and condition.

## Data Availability

All data is available with the corresponding author/first author

## Funding

Supported by a grant from the Intra-mural project of Era’s Lucknow Medical College and Hospital.

## Conflict of Interest

No conflict of interest, financial or other, exists.

## Notes

### Competing Interest Statement

The authors have declared no competing interest.

### Funding Statement

Supported by a grant from the Intra-mural project of Eras Lucknow Medical College and Hospital.

### Author Declarations

Study protocol was approved by institutional ethical committee of Eras Lucknow Medical College and Hospital.

